# Population-level risk factors related to measles case fatality: a conceptual framework based on expert consultation and literature review

**DOI:** 10.1101/2022.09.30.22280585

**Authors:** Alyssa N. Sbarra, Mark Jit, Jonathan F. Mosser, Matthew Ferrari, Felicity Cutts, Mark Papania, Katrina Kretsinger, Kevin A. McCarthy, Niket Thakkar, Katy A. M. Gaythorpe, Deepa Gamage, L. Kendall Krause, Emily Dansereau, Allison Portnoy

## Abstract

**Introduction:** A better understanding of population-level factors related to measles case fatality is needed to estimate measles mortality burden and impact of interventions such as vaccination. This study aimed to develop a conceptual framework of mechanisms associated with measles case fatality ratios (CFRs) and assess the scope of evidence available for related indicators in the published literature.

**Methods:** Using expert consultation, we developed a conceptual framework of mechanisms associated with measles CFR and identified population-level indicators potentially associated with each mechanism. We then conducted a literature review by searching PubMed on October 31, 2021 to determine and classify the scope of evidence for the indicators identified by expert consultation. Studies were included if they contained evidence of an association between an indicator and measles CFR and were excluded if they were from non-human studies or reported non-original data.

**Results:** Expert consultation identified five mechanisms in a conceptual framework of factors related to measles CFR: health system access and care-seeking behaviors, health system quality, measles control and epidemiology, nutritional status, and risk of secondary infection. Thirty-seven measurable population-level indicators were identified by expert consultation as proxies for the mechanisms. We identified 3772 studies for review and found 49 studies showing at least one significant association with CFR for 15 indicators and only non-significant associations for 5 indicators. Inadequate data were available to evaluate the remaining 17 indicators. A randomized controlled trial provided evidence for a relationship between vitamin A treatment and measles CFR. Average household size, educational attainment, first-dose coverage of measles-containing vaccine, human immunodeficiency virus prevalence, second-dose coverage of measles-containing vaccine, stunting prevalence, surrounding conflict, travel time to major city or settlement, travel time to nearest health care facility, under-five mortality rate, underweight prevalence, vitamin A deficiency prevalence, and wasting prevalence had quantitative observational level evidence. Level of health care available had qualitative evidence of a relationship with measles CFR.

**Conclusion:** To reduce uncertainty in measles CFR estimation, population-level factors representative of underlying mechanisms associated with CFR could be used. Our study used expert consultation and literature review to provide additional insights and a summary of the available evidence of these underlying mechanisms and indicators that could inform future estimations of measles CFR.

## Introduction

While measles mortality has decreased over the last several decades, an estimated 60,700 people died from measles in 2020.^1^ However in many settings, mortality is difficult to estimate through traditional measles case surveillance approaches alone due to challenges in cause-of-death attribution, weaknesses in vital registration systems, and variability in data completeness and quality in reporting cases and deaths.^2^ Instead, global stakeholders use models of measles mortality that require robust and dynamic estimates of measles case fatality ratios (CFR, i.e. proportion of cases with fatal outcome)^3^ to track progress towards eliminating measles deaths^4^ and to evaluate the impact of vaccination programs.^5^ Recently, an updated modeling approach has provided estimates of measles CFR by region, age group, and income-level for years 1990 through 2030.^6^ This foundation for producing dynamic CFR estimates is a critical advancement in estimating context- and intervention-specific measles mortality.^7^

There is evidence that various plausible risk factors contribute to systematically higher individual-level measles case fatality, such as nutritional or vaccination status, overcrowding at home, and overall health system access or quality.^8-10^ However, current surveillance systems do not systematically capture data on all possible risk factors for mortality. In places where accurate vital registration systems are not available, improved estimation of measles mortality burden, including the previously mentioned modeling approach, requires an understanding of case fatality risk factors. One approach is to evaluate evidence on potential CFR risk factors for which population-level data is consistently available, so that the most relevant population-level risk factors can be applied to estimates population-level CFR.

A clear framework of possible mechanisms related to CFR provides a means to organize the compiled evidence on risk factors associated with measles case fatality. Improved CFR estimates could help to assess health gains achieved by vaccination and other interventions such as nutrition supplementation, identify remaining gaps, understand likely drivers behind increased CFR, and support targeted efforts to reduce disease burden in particularly vulnerable communities. Therefore, we used expert consultation to develop a conceptual framework of mechanisms related to measles CFR and identify population-level indicators related to these underlying mechanisms. We used expert consultation because the underlying mechanisms related to measles case fatality are multifactorial, complex, and challenging to establish casual pathways of to describe systematic changes in CFR. Then, we conducted a literature review to assess the evidence of association between these indicators and case fatality.

## Methods

### Expert consultation

We consulted with a group of experts who are co-authors on this paper (Supplementary Information Section 1) to determine associative pathways that lead to either systematic increases or decreases in measles CFR. These pathways, referred to as “mechanisms” represent possible ways in which specific risk factors could be associated with measles CFR. We developed a conceptual framework relating each mechanism to measles case fatality.

To adequately represent these underlying mechanisms via population-level factors, we identified a list of 58 indicators typically available at the population level (Supplementary Information Section 2) that could be related to measles case fatality and together would be representative of these mechanisms. Following discussion, the group determined 42 of these possible indicators (Supplementary Information Section 3) to be most plausibly related to measles case fatality. From those, the group determined a list of indicators for further investigation with at least one vote for their inclusion (Supplementary Information Section 4); through this process, exclusive breastfeeding and sanitation quality indicators were removed. Age, measles incidence / attack rate, and outbreak settings have complex interactions with each other and other indicators as well, and as such, were determined to be fundamental in any consideration of measles mortality without requiring further investigation in the literature.^6,10^ This yielded 37 indicators for additional investigation. Each indicator was then assigned to represent an underlying mechanism following discussion of the expert group.

### Literature review

To assess the level of evidence of association between measles case fatality and the final list of identified indicators, we conducted a review of the available literature (Supplementary Information Section 5). We searched the PubMed database from January 1, 1980 to October 31, 2021 for any article with the following search terms:

> (indicator-specific search terms)
>
> AND “measles”
>
> AND (“case fatality” OR “CFR” OR “fatality” OR “mortality” OR “morbid*” OR “comorbid*” OR
>
> “sever*” OR “complicat*” OR “risk” OR “secondary outcome” OR “death”)

A full list of indicator-specific search terms can be found in Supplementary Information Section 6. Articles were included if they contained information on an association between measles case fatality or acute mortality and the indicator of interest among any age or setting. Articles were excluded if they were from non-human studies or reporting on non-original data. Following the search, each indicator was assigned to one of the following categories: indicator has at least one published randomized controlled trial supporting a significant relationship with CFR, indicator has at least one published quantitative observational study supporting a significant association with CFR, indicator has at least one published qualitative study supporting an association with CFR, indicator has published evidence of a non-significant relationship between indicator and CFR, and indicator had no published evidence investigating the relationship with CFR, depending on the highest quality evidence found.

## Results

### Conceptual framework

Five underlying mechanisms were identified by the expert group: health system access and care-seeking behaviors, health system quality, measles control and epidemiology, nutritional status, and risk of secondary infection. Each mechanism was hypothesized by the expert group to have a direct association to either systematic increases or decreases in measles CFR, as well as interdependently with one another (Figure 1). For example, risk of secondary infection would be directly associated with measles CFR, but would also be associated with nutritional status, which would also be directly associated with measles CFR. Each indicator outlined above was assigned a primary mechanism (Table 1) to ensure each mechanism was adequately represented by the grouping of indicators assigned to it (e.g., human immunodeficiency virus, or HIV, prevalence was assigned to the “risk of secondary infection” mechanism).

**Table 1.** Mechanisms impacting measles CFR with related hypothesized indicators

| Mechanisms | Indicators |
| --- | --- |
| Health system access and care-seeking behaviors | • Educational attainment<br>• Percent living in urban settings<br>• Surrounding conflict<br>• Time to care seeking<br>• Travel time to major city or settlement<br>• Travel time to nearest health care facility |
| Health system quality | • Access to intensive care unit<br>• Health expenditure per capita<br>• Level of health care available<br>• Under-five mortality rate |
| Measles control and epidemiology | • First-dose coverage of measles-containing vaccine (MCV1)<br>• Maternal antibody dynamics<br>• Maternal measles vaccination coverage<br>• Second-dose coverage of measles-containing vaccine (MCV2)<br>• Vaccine coverage equity<br>• Vaccination efficacy<br>• Vaccination schedule<br>• Vitamin A treatment |
| Nutritional status | • Stunting prevalence<br>• Underweight prevalence<br>• Vitamin A deficiency prevalence<br>• Vitamin A supplementation<br>• Wasting prevalence |
| Risk of secondary infection | • Ambient air pollution<br>• Antibiotic use for measles-related pneumonia<br>• Average household size<br>• De-worming frequency<br>• Diarrheal disease prevalence<br>• Human immunodeficiency virus (HIV) prevalence<br>• Human immunodeficiency virus (HIV) treatment / antiretroviral therapy (ART) prevalence<br>• Malaria prevalence<br>• Lower respiratory infection prevalence<br>• Oral rehydration solution for measles-related diarrhea<br>• Pneumococcal conjugate vaccination coverage<br>• Population density<br>• Pre-term birth prevalence<br>• Total fertility rate |

**Figure 1.**
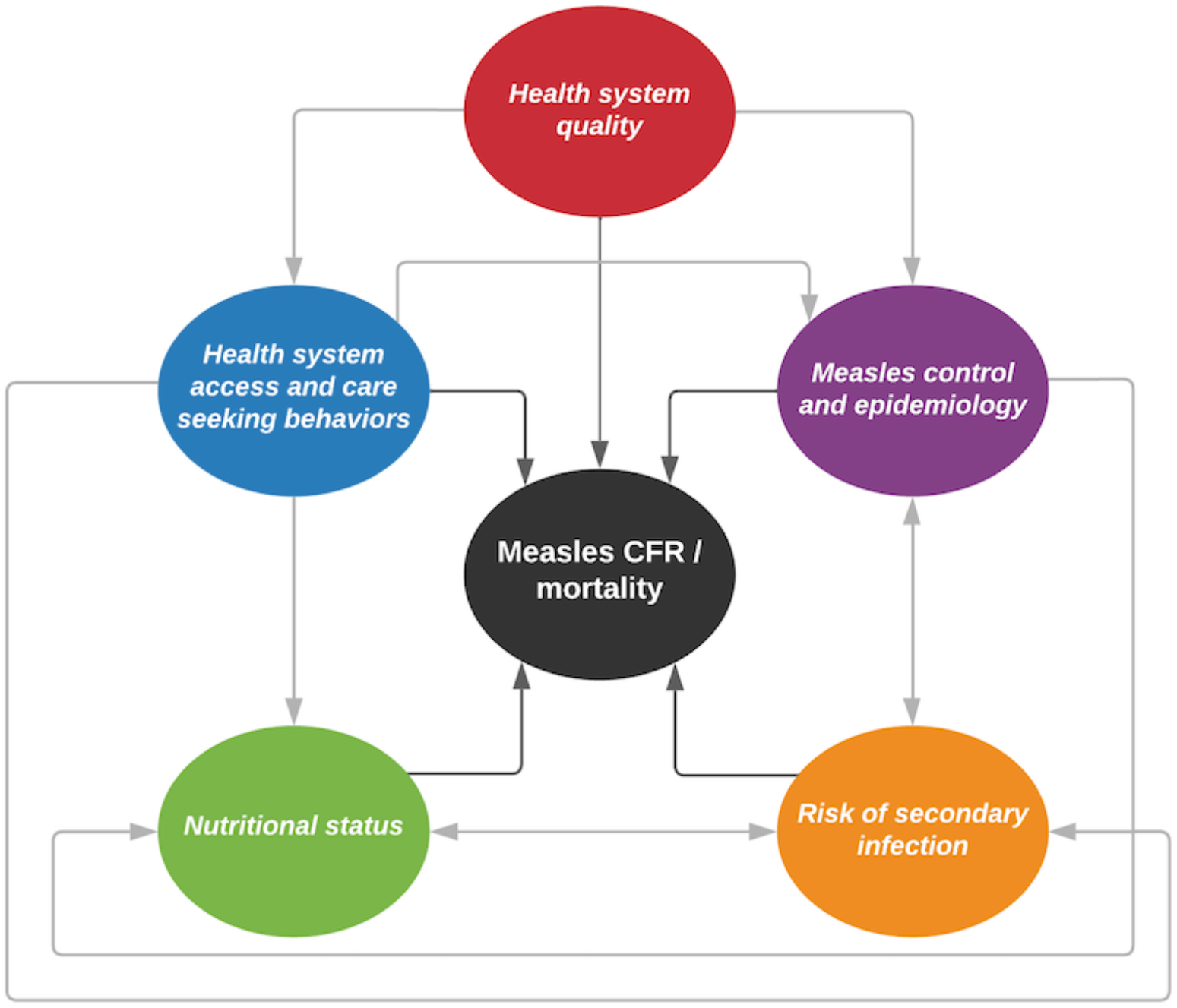
Conceptual framework of mechanisms related to measles case fatality rates (CFRs). Each mechanism is represented by a colored circle. Dark grey arrows show direct relationship with measles CFR and light grey arrows show relationships between the mechanisms.

### Literature review

The search yielded 3772 articles; full text was reviewed for 857 of these articles (Figure 2). Each indicator was classified depending on availability of evidence (Table 2). There was one indicator with at least one published randomized controlled trial supporting a significant relationship with CFR, 13 indicators with at least one published quantitative observational study supporting a significant association with CFR, one indicator with at least one published qualitative study supporting an association with CFR, five indicators with published evidence of a non-significant relationship between indicator and CFR, and 17 indicators with no published evidence investigating a relationship with CFR. For the 49 studies in which there was evidence of an association for an indicator with measles case fatality, findings are outlined below.

**Table 2.** Available evidence of relationship between indicators and measles CFR.

| Published literature includes randomized controlled trial with significant relationship | Published literature supports significant observational association | Published literature supports qualitative association | Published literature with non-significant evidence | No evidence found in published literature |
| --- | --- | --- | --- | --- |
| Vitamin A treatment | Average household size<br>Educational attainment<br>First-dose coverage of measles-containing vaccine (MCV1)<br>Human immunodeficiency virus (HIV) prevalence<br>Second-dose coverage of measles-containing vaccine (MCV2)<br>Stunting prevalence<br>Surrounding conflict<br>Travel time to major city or settlement<br>Travel time to nearest health care facility<br>Under-five mortality rate<br>Underweight prevalence<br>Vitamin A deficiency prevalence<br>General malnutrition (surrogate for wasting prevalence) | Level of health care available | Antibiotic use for measles-related pneumonia<br>Malaria prevalence<br>Percent living in urban settings<br>Pneumococcal conjugate vaccination coverage<br>Vitamin A supplementation | Access to intensive care unit<br>Ambient air pollution<br>De-worming frequency<br>Diarrheal disease prevalence<br>Health expenditure per capita<br>Human immunodeficiency virus (HIV) treatment / antiretroviral therapy (ART) prevalence<br>Lower respiratory infection prevalence<br>Maternal antibody dynamics<br>Maternal measles vaccination coverage<br>Oral rehydration solution for measles-related diarrhea<br>Population density<br>Pre-term birth prevalence*<br>Time to care seeking<br>Total fertility rate<br>Vaccine coverage equity<br>Vaccination efficacy<br>Vaccination schedule |

Results from literature review are summarized per indicator into one of five categories describing the level of evidence available from published literature.
\* Evidence was found for an association but excluded for only having one study with significant evidence which had a small sample size across entire body of. evidence for this indicator.

**Figure 2.**
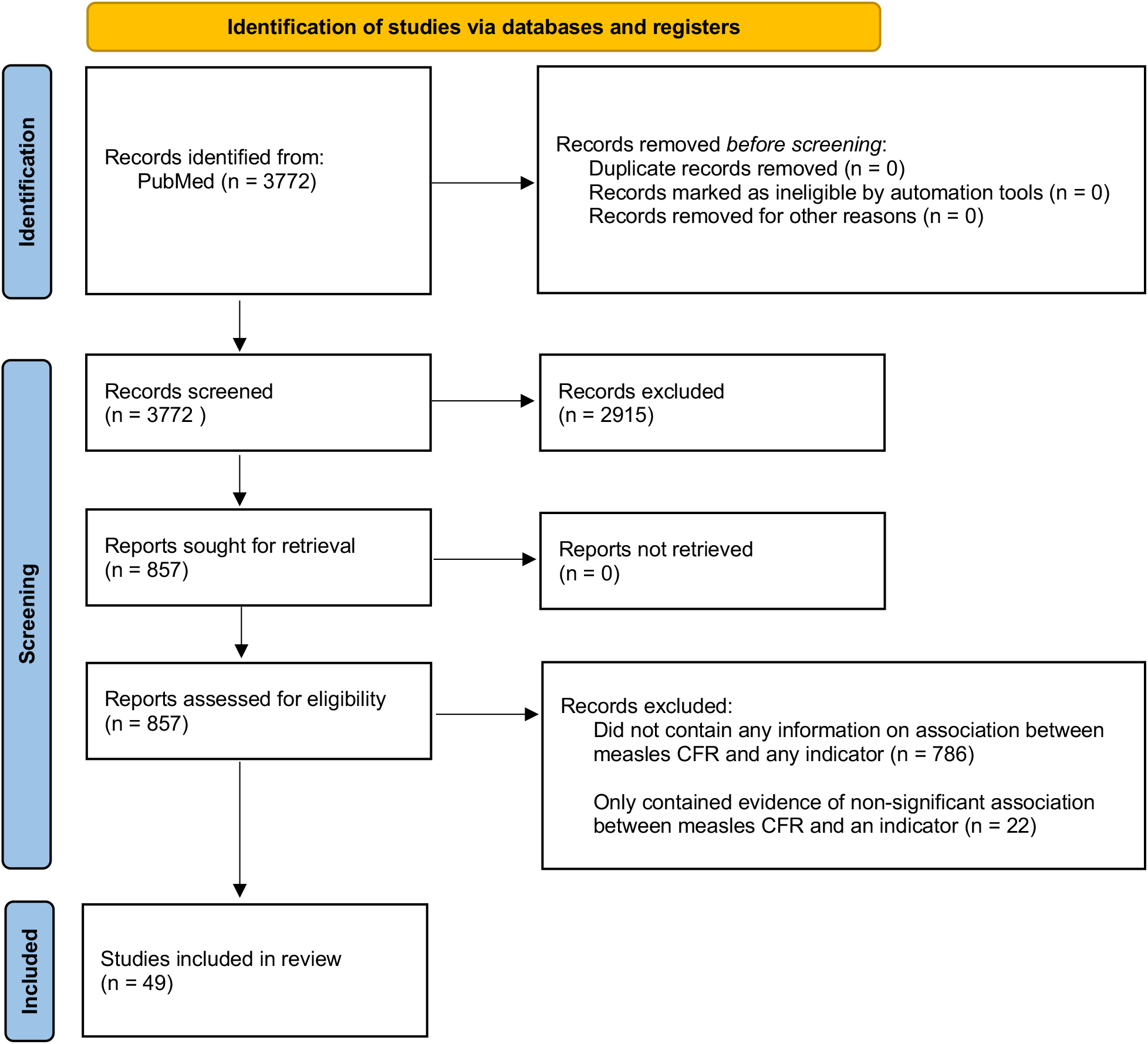
PRISMA diagram of literature review sources. For the completed literature review, number of studies at each stage of the review is shown. Studies included in review were those to have contained significant evidence of an association with an indicator and measles CFR.

Among these 49 studies, the following 26 countries were represented: Afghanistan, Bangladesh, Chad, Costa Rica, Democratic Republic of the Congo, Ethiopia, France, Ghana, Guinea-Bissau, India, Indonesia, Kenya, Malawi, Mexico, Mongolia, Nepal, Niger, Nigeria, Pakistan, Papua New Guinea, Senegal, South Africa, Sweden, Tanzania, Turkey, and Zambia. There were 9 studies published from 1980–1989, 14 from 1990–1999, 14 from 2000–2009, 7 from 2010–2019, and 5 from 2020–2021. All studies with significant and non-significant evidence of an association between an indicator and measles CFR are presented in Supplementary Information Sections 7–8.

#### Health system access & care-seeking behaviors

##### Educational attainment

Bhuiya and colleagues^11^ showed in 1980 that Bangladeshi children with measles whose mothers had no education had increased odds of death (odds ratio (OR): 2.11 [95% confidence interval (CI): 1.06–4.19]) compared to those with mothers having “some” education. In a case-control study among deaths and non-fatal cases in Bangladesh from 1982 to 1984, Clemens and colleagues^12^ estimated increased odds (OR: 1.32 [1.07–1.63]) of death when the head of household had no (versus any) education. Similarly, they found increased odds (OR: 1.72 [1.36–2.19]) of death when mothers had no (versus any) education. Ndikuyeze and colleagues^13^ noted there were higher CFRs among children living in Chad from 1988 to 1992 with mothers who were “less well educated”; no data were shown. In Zambia from 1998–2003, Moss and colleagues^14^ showed children hospitalized with measles whose maternal education was less than or equal to eight years had an increased relative risk of death (relative risk (RR): 2.15 [1.11–4.17]) compared to children whose mothers had more than eight years of education. Murhekar and colleagues^15^ showed that children with measles with illiterate heads of households living in India in 2012 had an increased relative risk (RR: 6.23 [1.48–26.21]) of death compared to those having a parent with primary education or above.

##### Surrounding conflict

Salama and colleagues^16^ noted that in 2000 in Ethiopia, famine, exacerbated by conflict, was associated with many deaths related to measles, and without relief interventions, measles mortality would have been greater. Joshi and colleagues^17^ showed that, in Nepal in 2004, children with measles living in locations with critical insecurity levels had increased odds of death (OR: 15.8 [3.4–73.4]) compared to children living in locations with moderate insecurity levels. Additionally, Meteke and colleagues^18^ noted conflict settings can lead to infectious disease outbreaks; no data were shown. Moss and colleagues^19^ noted that measles is a major cause of death among internally displaced and refugee persons as well as very high rates of CFRs among various emergency situations; no data were shown.

##### Travel time to nearest health facility / major city or settlement

In 2013 in the Democratic Republic of the Congo, Gignoux and colleagues^20^ showed children with measles who lived more than 30 kilometers from a hospital had an increased relative risk (RR: 2.2 [1.0–4.7]) of death compared to children with measles who lived 30 or fewer kilometers away from a hospital. From 2015-16 in Mongolia, Lee and colleagues^21^ noted that children with measles who lived outside of Ulaanbaatar City had an increased relative risk (RR: 1.9 [1.3–2.8]) of death compared to children with measles who lived within Ulaanbaatar City.

##### Indicators without supporting evidence

For percent living in urban settings, published evidence of a non-significant relationship with CFR was found. There was no published evidence examining the association between CFR and time to care seeking.

#### Health system quality

##### Level of health care available

Rey and colleagues^22^ noted, among measles cases in France from 1970–1979, there was a significant decrease in mortality throughout the study period that the authors suggested was most likely attributable to general improvements in health care availability; no data were shown.

##### Under-five mortality rate

Multiple studies^16,23-26^ noted the temporal correlation and overall trend between decreasing under-five mortality and measles mortality in various years and settings.

##### Indicators without supporting evidence

There was no published evidence examining the association between CFR and access to intensive care units or health expenditure per capita.

#### Nutritional status

##### Malnutrition

Joshi and colleagues^17^ showed that in 2004 in Nepal, persons with measles who were stunted had an increased relative risk (RR: 5.34 [2.31–12.36]) of death compared with persons who were not stunted. Among persons hospitalized with measles from 2013 to 2017 in Pakistan, Aurangzeb and colleagues^27^ estimated that there was increased odds (OR: 6.8 [3.24–14.26]) of death among those who were stunted compared to those who were not stunted.

Barclay and colleagues^28^ showed that, among persons with measles from 1982 to 1983 in Tanzania, there was an increased relative risk (RR: 3.94 [1.69–9.21]) of death among persons with a weight-for-age less than 60%. In Ghana from 1989 to 1991, Dollimore and colleagues^29^ described increased odds (OR: 2.5 [1.3–5.1], adjusted for age, sex, vaccination status, paternal education, and wet versus dry season) of death among children with measles who were two or more standard deviations below their weight-for-age z-score compared to those that were not. Among patients hospitalized with measles from 2000 to 2004 in Nigeria, Lagunju and colleagues^30^ showed that underweight persons had an increased relative risk (RR: 2.23 [1.17–4.26]) of death compared to those who were not underweight. Additionally, Ahmed and colleagues^31^ estimated that in 2002 to 2005 in Nigeria, being underweight was associated with increased measles case fatality (chi-squared p=0.01). In South Africa from 2009 to 2010, le Roux and colleagues^32^ showed that there was an association between overall weight-for-age and measles case fatality. Also, among persons hospitalized with measles from 2013 to 2017 in Pakistan, Aurangzeb and colleagues^27^ noted that there was increased odds (OR: 2.93 [1.44–5.93]) of death among measles cases who were underweight compared to those who were not underweight. Coetzee and colleagues^33^ showed that among pediatric hospitalized measles patients in an intensive care unit in South Africa in 2014, underweight persons had an increased relative risk (RR: 2.77 [1.38–5.55]) of death compared to persons who were not underweight.

In addition to the biometric indicators of stunting and underweight identified by the expert group, multiple studies reported on the association between non-specific malnutrition and measles case-fatality; as such, the evidence associating non-specific malnutrition with measles case fatality are presented here. From 1973 to 1982 in Ghana, Commey and colleagues^34^ showed that among measles hospitalizations for malnourished children, there was an increased relative risk (RR: 2.02 [1.63–2.51]) of death compared to measles hospitalizations among children who were not malnourished. Avila-Figueroa and colleagues^35^ showed that children hospitalized with measles from 1976 to 1989 in Mexico had an increased relative risk (RR: 2.47 [1.1–5.52]) of death if they were malnourished compared to those who were not malnourished. Samsi and colleagues^36^ showed that hospitalized measles patients in Indonesia from 1982 to 1986 who were malnourished had an increased relative risk of death (RR: 2.48 [1.4–4.39]) compared to hospitalized measles patients who were not malnourished. Among hospitalized measles patients in Kenya from 1982 to 1985, Alwar and colleagues^37^ showed that those who were malnourished had an increased relative risk (RR: 3.77 [1.85–7.66]) of death compared to those who were not malnourished. Choudhry and colleagues^38^ noted that in Afghanistan from 1983 to 1985, hospitalized measles cases with malnutrition had an increased relative risk (RR: 14.66 [5.46–39.36]) of death compared to patients without malnutrition. Madhulika and colleagues^39^ noted that in India in 1991 measles cases that were malnourished experienced increased case fatality compared to those that were not malnourished (Chi-squared p=0.0156). In hospitalized measles patients from 1994 to 2004 in Nigeria, Fetuga and colleagues^40^ estimated that cases who were malnourished had an increased relative risk (RR: 7.33 [1.62– 33.16]) of death compared to patients who were not malnourished. Moss and colleagues^19,41^ also noted an association between malnutrition and case fatality; no data were shown. The expert group identified wasting as a potential biometric indicator related to CFR, however no evidence directly examining this relationship was found. However, because of the strong preponderance of evidence supporting an association between general malnutrition and CFR, wasting was included having observational-level associative evidence.

##### Vitamin A deficiency prevalence

Among hospitalized measles patients in Malawi from 1992 to 1993, Courtright and colleagues^42^ showed that there was an increased relative risk (RR: 4.00 [1.21–13.33]) of death for patients with vitamin A abnormalities compared to patients without vitamin A abnormalities. Moss and colleagues^19^ noted that there is a high risk of measles case fatality for those with underlying vitamin A deficiencies; no data were shown. Additionally, Nojilana and colleagues^43^ reported on the increased relative risk (RR: 1.86 [1.32– 2.59]) of vitamin A deficiency on measles case fatality.

##### Vitamin A supplementation

While evidence was collected from various studies on vitamin A supplementation, a recent meta-analysis^44^ pooling data from 43 trials in 18 settings between 1976–2010 found no effect of vitamin A supplementation on measles case fatality. We, therefore, chose to exclude this indicator.

#### Risk of secondary infection

##### Average household size

Burström and colleagues^45^ showed that from 1885 to 1910 in Sweden there was an increased relative risk of death among children with measles who had siblings (RR: 2.9 [1.6–5.4]) compared to those who did not, as well as for children with measles with a household size of more than four persons (RR: 1.9 [1.3–2.8]) compared to those with household sizes with three or less people; they also showed other significant univariate associations. Aaby and colleagues^46^ noted that among children in Guinea-Bissau in 1979 there was an association between increased case fatality among “other” types of households relative to those that were monogamous (Chi-squared p<0.01). Also, Aaby and colleagues^47^ showed that among children with measles in Guinea-Bissau in 1979, those living in homes with more than four children had an increased relative risk (RR: 1.9 [1.2 – 3.0]) of death compared to those living in homes with four or fewer children. Nandy and colleagues^48^ noted that among persons with measles in Niger in 2003, those living in a household with eight or more persons had an increased relative risk (RR: 1.82 [1.22 – 2.71]) of death compared to those living with fewer than eight persons.

##### HIV prevalence

Among hospitalized measles patients from 1993 to 1995 in Zambia, Oshitani and colleagues^49^ found that patients with HIV had an increased relative risk (RR: 3.35 [1.95–5.76]) of death compared to patients without HIV. Jeena and colleagues^50^ showed that hospitalized measles patients from 1994 to 1996 in South Africa who were HIV positive had a substantially increased relative risk (RR: 129.62 [40.12–412.64]) of death compared to patients who were HIV negative. Moss and colleagues^14^ demonstrated that from 1998–2003 in Zambia, hospitalized measles patients with HIV had an increased relative risk (RR: 2.95 [1.83–4.74]) of death when compared to other hospitalized measles patients without HIV. Also, in an outbreak with 552 cases from 2009 to 2010 in South Africa, le Roux and colleagues^32^ estimated that after adjusting for age and weight-for-age, persons with HIV had increased odds (OR: 7.55 [2.27–25.12]) of death compared to persons without HIV. Coetzee and colleagues^33^ also showed that among pediatric hospitalized measles patients in an intensive care unit in South Africa in 2014, persons with HIV had an increased relative risk (RR: 2.29 [1.24–4.20]) of death compared to persons without HIV.

##### Indicators without supporting evidence

Published evidence of non-significant relationships between each indicator and CFR were found for the following indicators: antibiotic use for measles-related pneumonia, malaria prevalence, and pneumococcal conjugate vaccination coverage. There was no published evidence examining the association with CFR and the following indicators: ambient air pollution, human immunodeficiency virus (HIV) treatment / antiretroviral therapy (ART) prevalence, de-worming frequency, oral rehydration solution for measles-related diarrhea, population density, and total fertility rate. For the association between pre-term birth prevalence and increased measles case fatality, evidence of an association^51^ was found but excluded for only having one study with significant evidence which had a small sample size (N=57). Several studies suggested a significant relationship between measles case fatality and diarrheal disease or lower respiratory infection. In these studies, however, it was not possible to distinguish whether these data reflected secondary infections or measles-virus related symptoms. After discussions with the expert group, we excluded these studies and these indicators.

#### Measles control and epidemiology

##### First-dose coverage of measles-containing vaccine (MCV1)

Nayir and colleagues^52^ noted a temporal trend relating measles vaccination and declining measles mortality rates in Turkey from 1970 to 2017. In Bangladesh from 1982 to 1985, Aaby and colleagues^53^ showed a protective effect of measles vaccination on measles deaths (vaccine efficacy against measles death: 95% [79% - 99%]). Samb and colleagues^54^ estimated that vaccinated measles cases in Senegal from 1983 to 1990 had lower case fatality rates than unvaccinated cases (p=0.038). Oshitani and colleagues^55^ showed that in Zambia from 1992 to 1993, children with measles who had at least one dose of any measles-containing vaccine (MCV) had a decreased relative risk (RR: 0.4 [0.19–0.83]) of death compared to unvaccinated children with measles. Fetuga and colleagues^40^ noted an association between measles vaccination and case fatality in Nigeria from 1994 to 2004 (Fisher’s exact p=0.033). Dollimore and colleagues^29^ noted that among measles cases from 1998 to 1999 in Ghana, those who were unvaccinated had an increased relative risk (RR: 1.72 [1.04–2.84]) of death compared to those who were previously vaccinated with at least one dose of any MCV. Mgone and colleagues^56^ noted an inverse association between measles vaccination and case fatality in Papua New Guinea in 1999 (chi-squared p=0.0423) among patients hospitalized with measles.

Among hospitalized measles patients from 2003 to 2004 in Pakistan, Aurangzeb and colleagues^57^ showed that there were increased odds (OR: 8.40 [1.00–71.84]) of death among children who were previously unvaccinated compared to those with at least one dose of MCV. Among persons with measles in Nepal in 2004, Joshi and colleagues^17^ showed that there was an increased relative risk (RR: 3.7 [2.0–6.7]) of death among unvaccinated cases compared to those who had at least one dose of any MCV. From 2007 to 2016 in Ethiopia, Gutu and colleagues^58^ showed that unvaccinated measles cases had increased odds (OR: 1.55 [1.14–2.11]) of death compared to cases with a previous vaccination history with at least one dose of any MCV. Gignoux and colleagues^20^, in a 2013 outbreak in the Democratic Republic of the Congo, noted a decreased relative risk (RR: 0.3 [0.1–0.9]) of death among children previously vaccinated with only one dose of MCV compared to those that were previously unvaccinated. Moss and colleagues^19,41^ have also noted an association between measles vaccination status and case fatality; no data were shown.

##### Second-dose coverage of measles-containing vaccine (MCV2)

During an outbreak in the Democratic Republic of the Congo in 2013, Gignoux and colleagues^20^ noted a decreased relative risk (RR: 0.2 [0.1–0.3]) of death among children with measles who were previously vaccinated with at least two doses of MCV (compared to those that were previously unvaccinated or had only received one dose). Aurangzeb and colleagues^27^ showed that in Pakistan from 2013 to 2017, children with measles that were previously unvaccinated had an increased relative risk (RR: 7.0 [2.03–24.01]) of death compared to those who had received at least two doses of any MCV; additionally, children with measles that had previously only had one dose had an increased relative risk (RR: 5.73 [1.49–22.07]) of death compared to those who had received at least two doses of any MCV.

##### Vitamin A treatment

In a randomized trial in 1987 in South Africa, Hussey and colleagues^59^ showed there were decreased odds (OR: 0.21 [0.05–0.94]) of death among hospitalized measles patients who had received vitamin A treatment compared to those who had not. In another randomized trial from 1989–1990 in South Africa, Hussey and colleagues^60^ concluded there were decreased odds (OR: 0.36 [0.18–0.70]) of death among hospitalized measles patients who were treated with vitamin A therapy compared to those who were not. Joshi and colleagues^17^ showed that in 2004 in Nepal measles cases that did not receive vitamin A treatment had an increased relative risk (RR: 3.09 [1.69–5.67]) of death compared to cases that did receive vitamin A treatment. Murhekar and colleagues^15^ showed that in India in 2012, among measles cases who received vitamin A treatment, there was a decreased relative risk (RR: 0.14 [0.03–0.61]) of death compared to those who did not receive treatment. In a measles outbreak in 2017 in India, Dzeyie and colleagues^61^ showed vitamin A treatment was associated with decreased measles case fatality (chi-squared p=0.0351).

##### Indicators without supporting evidence

We did not find any studies examining the relationship between CFR and the following indicators: maternal measles vaccination coverage, maternal antibody dynamics, vaccination efficacy, vaccination schedule, and vaccine coverage equity.

## Discussion

Our conceptual framework of mechanisms related to measles CFR, based on expert consultation, and literature review of indicators associated with these mechanisms strengthen the understanding of measles CFR and mortality estimation. We categorized potential risk factors for measles CFR into five mechanisms related to either systematic increases or decreases in measles CFR and searched for evidence of an association with measles CFR across 37 population-level indicators that are representative of these mechanisms. Among indicators included in our search, 15 indicators had evidence of an association with measles CFR.

Overall, 26 countries were represented in 49 studies published from 1983 to 2021 that included quantitative or qualitative evidence of an association with CFR. Most locations were from low- or middle-income countries. Relative to other mechanisms, nutritional status had the greatest number of studies available across its group indicators. Only one indicator, vitamin A supplementation, had a previously conducted systematic review with meta-analysis pooling results across studies. For multiple indicators, results of various studies showed both statistically significant and non-significant associations with CFR. In the absence of a meta-analysis, we considered indicators with literature presenting evidence of a statistically significant association with CFR and literature presenting evidence of a non-significant association to be plausibly associated with CFR if there was at least one study with significant evidence.

Most indicators could likely be classified under multiple underlying mechanisms. For example, MCV1 coverage could be related to measles control and epidemiology, but also to general health system access, health system quality, and risk of secondary infection. However, because indicators were used to represent mechanisms at large, the assignment of each indicator to a single mechanism for illustrative purposes did not influence the determination as to whether an association with CFR existed.

While they may be associated with measles CFR, we identified 17 indicators for which no evidence had any significant association. For some indicators, the type of data available did not allow us to reliably assess the association with CFR, despite the availability of published evidence. For lower respiratory infection and diarrheal disease prevalence, high rates of community prevalence of related pathogens may theoretically increase the risk of secondary infection in measles cases and subsequently increase case fatality. There was substantial evidence to suggest that the development of pneumonia or diarrhea following acute measles infection was associated with increased case fatality. However, it was not possible to distinguish whether the development of pneumonia or diarrhea represented progression of primary measles or reflected secondary infection with an additional pathogen. Without routine specimen testing when additional clinical symptoms arise, we are unable to distinguish whether these are population-level factors related to increased measles CFR or markers of disease severity. Thus, it was not possible to determine the nature of these specific associations.

Given the heterogeneity of underlying studies, including study design and sample size, we were unable to perform any quantitative synthesis to combine the evidence found in the published literature.

Additionally, the objective of our review was to generate supportive evidence for the conceptual framework and related indicators via identifying any evidence suggesting an association with measles CFR rather than generating a single effect size per indicator.

We assessed only associations with acute measles case fatality as our end point. However, it is known that since health facilities experience higher patient loads^62^ and secondary measles cases present with increased severity compared to primary cases^47,63^, increases in measles incidence are associated with increased measles CFR (such as in an outbreak setting^10^). If this association is causal, then anything that increases measles incidence could also plausibly increase measles CFR, but we did not examine these relationships in this work.

We did not explicitly re-examine the relationship between age or measles incidence, given their known importance regarding case fatality. It has been shown previously that as age increases, CFR decreases.^6,10^ These patterns likely reflect a variety of complex relationships between age, maturation of immune responses to infection, and age-dependencies in other risk factors for CFR. In young infants, maternal antibodies are likely to provide some protection both against infection and case fatality, though the presence and duration of this protection depends on maternal immunity rates, gestational age, and underlying nutritional status, among other additional factors.^64^ As maternal immunity wanes, young children may be particularly vulnerable to measles infection and case fatality, until they receive measles vaccination (typically between 9–12 months). The complex interplay between maternal immunity, measles epidemiology, and vaccination (as well as possible age-related confounders) complicates interpretation of reported CFRs—especially among age groups representing these youngest children— and warrants particular attention when developing measles control strategies. Complex relationships between other risk factors for measles CFR—such as between MCV1 coverage and HIV prevalence—can also contribute to differences in measles CFR by age and may vary from setting to setting. Children born to HIV-positive mothers are likely to have fewer maternal antibodies^65^ as well as a lower probability of sustained seroconversion following measles vaccination.^66^ Persons living with HIV are more likely to both acquire and subsequently die from measles, making them a particularly important community to consider when estimating measles CFR.^67^ More robust data needs to be collected to better understand and account for these interdependent relationships between age, measles incidence, MCV coverage, and HIV prevalence. Additionally, since the relationship between age and measles CFR is so strong, modeling efforts to understand measles mortality should ideally account for the underlying age pattern in both measles cases and CFRs.

This work has several limitations. First, the evidence presented in this study is heterogeneous and includes both population-level and individual-level relationships. Next, we did not identify the reasons for studies showing non-significant associations, such as having an underpowered sample size to assess significance. Additionally, due to limitations in available data, we were unable to assess causality of associations between indicators and CFR. For example, as with nutritional status, few studies had information on anthropometry prior to measles onset, and since measles infection commonly leads to weight loss, reverse causality cannot always be excluded. Nonetheless, some large prospective studies confirmed increased CFR in malnourished children. Additionally, we did not consider evidence specific to populations that are at particularly high risk of measles infection and mortality, such as refugees or internally displaced persons. While these subgroups of the overall population are likely at higher risk of both measles infection and mortality given underlying concerns related to access to health services and other increased risk of infection, there is a scarcity of population-specific indicators and underlying data regarding measles CFR. Although data in these populations are likely challenging to collect, we support investigation of these critical questions to better understand how to assess burden among these high-risk groups. Additionally, we considered only acute fatality from a measles case (i.e., within the first 28 days). Additional consideration should be given to which indicators and mechanisms contribute to longer term impacts of measles on overall mortality.^68^ Finally, several studies did not provide information on either the proportion of cases with laboratory confirmation or the underlying definition for a measles case, which reduces the overall quality of evidence presented in these particular studies.

Overall, this study addresses some of the knowledge gaps around factors influencing measles CFR and, moreover, may be valuable for decision making and programmatic targeting among disease control programs. More work as well as primary data collection are needed to continue to expand what is known about these associations, to close important knowledge gaps, and to better estimate measles CFR across settings and populations.

## Data Availability

All data produced in the present work are contained in the manuscript.

## Acknowledgements

This study was funded by the Bill & Melinda Gates Foundation. We thank Natasha Crowcroft and Patrick O’Connor for their valuable and thoughtful comments.

## Contributions

ANS, MJ, JFM, and AP conceived and planned the study. MJ, JFM, MF, FC, MP, KK, KAM, NT, KAMG, DG, LKK, and ED participated as members of the expert group, which was coordinated by ANS and AP and supported by MJ and JFM. ANS and AP performed the literature review (i.e., screened, extracted, and synthesized data). ANS wrote the first draft of the manuscript and all authors contributed to subsequent revisions and approved the final manuscript as submitted. All authors provided intellectual input into aspects of this study.

