## Supplementary Information for "Population-level risk factors related to measles case fatality: a conceptual framework based on expert consultation and literature review"

### Section 1. Activities of the Expert Working Group

The Working Group was established with the intent to gain feedback throughout the development of a conceptual framework as related to measles CFR. Members of the Working Group included:

- Natasha Crowcroft (WHO)
- Felicity Cutts (LSHTM)
- Emily Dansereau (BMGF)
- Matthew Ferrari (Penn State)
- Deepa Gamage (WHO)
- Katy Gaythorpe (VIMC)
- Kendall Krause (BMGF)
- Katrina Kretsinger (WHO / CDC)
- Kevin McCarthy (IDM / BMGF)
- Mark Papania (CDC)
- Niket Thakkar (IDM / BMGF)

The overall objectives for convening this Working Group, related to indicator investigation, were as follows:

1. Determine all possible indicators (and proxy metrics, as needed) as related to measles CFR
2. Determine relative order and group of available indicator importance
3. Provide guidance and recommendation on targeted literature reviews
4. Approve final indicator list and conceptual framework

Each session of the Working Group was conducted online, via Zoom. Activities for each Session are outlined below.

#### Session 1

*Objectives*

1. *Discuss a full list of possible covariates to explore, including proxies*
2. *Determine overall importance of each covariate candidate*

Working Group members brainstormed a comprehensive, full list of possible population-level indicators related to measles CFR. These were not to include those describing special populations, such as internally displaced persons and refugees, as the underlying available CFR data does not include adequate information on these groups. The full list of indicators generated by the Working Group are defined in Supplementary Information Section 2. Working Group members voted for each indicator that they thought had an important relationship with measles CFR; members could “up-vote” or “down-vote” for each. Indicators with no more than 2 down-votes were considered for further inclusion. This list can be found in Supplementary Information Section 3.

#### Session 2

*Objectives:*

1. *Anonymously rank indicators to determine relative importance*
2. *Determine indicator candidates further worth investigation*

Working Group members ranked indicators in order of importance to consider relative to one another. Each ranked position from 1 to 42 was assigned each corresponding weight. Age was removed from this process. Overall indicator rank was determined by average weight across responses, shown in Supplementary Section 4.

Members asserted, with at least one verbal yes, their desire for the inclusion of all indicators for further analysis. Members suggested considering mechanisms that might impact measles mortality or case fatality so it could be ensured that remaining indicators adequately captured the underlying components of these possible mechanisms.

#### Session 3

*Objectives*

1. *Review mechanistic groups and indicators per group*
2. *Review protocol for literature review and dataset investigation*

Members reviewed the following mechanistic groups and indicators corresponding with each group. The groups and related covariates are described in Table 1 (main text).

Members confirmed the inclusion of all indicators other than sanitation quality and the following protocol for literature review and data analysis:

1. Search for and review any available literature (systematic literature review)
2. Search for and review any available population level data (database search)
3. Categorize into following groups:
    a. Published literature supporting causal relationship and population-level data
    b. Published literature supporting observational relationship and population-level data
    c. Published literature with supporting qualitative evidence and population-level data
    d. No literature published, but population-level data available
    e. No literature published and population-level data is untrustworthy, contains missingness, or is otherwise unsuitable
4. Follow-up with Working Group to share indicator categories
5. Framework development

#### Session 4

*Objectives*

1. *Provide feedback on proposed conceptual framework of mechanistic groups*
2. *Review results from literature review and dataset investigation*
3. *Provide specific recommendation for areas in literature with ambiguous results*

From a systematic review of the literature, each covariate was classified in Table 2 (main text).

### Section 2. Full list of identified indicators potentially related to measles case fatality identified by Expert Working Group

- Access to intensive care unit (ICU)
- Age
- Ambient air pollution
- Antibiotic use for measles-related pneumonia
- Asthma prevalence
- Autoimmune condition prevalence
- Average household size
- Bacille Calmette-Guérin vaccination coverage
- Breastfeeding prevalence
- Cancer prevalence
- De-worming frequency
- Diarrheal disease prevalence
- Diphtheria- tetanus- pertussis (DTP) vaccination coverage
- Educational attainment
- First-dose coverage of measles-containing vaccine (MCV1)
- Health expenditure per capita
- *Haemophilus influenzae* type B (Hib) vaccination coverage
- Human immunodeficiency virus (HIV) prevalence
- Human immunodeficiency virus (HIV) treatment / antiretroviral therapy (ART) prevalence
- Household air pollution
- Level of health care available
- Lower respiratory infection (LRI) prevalence
- Malaria prevalence
- Maternal antibody dynamics
- Maternal measles vaccination coverage
- Maternal smoking prevalence
- Measles attack rate / incidence
- Meningococcal serogroup A vaccination coverage
- Oral rehydration treatment or solution (ORT/S) for measles-related diarrhea
- Outbreak susceptibility
- Overweight prevalence
- Pneumococcal conjugate vaccination (PCV) coverage
- Polio vaccination coverage
- Pre-term birth prevalence
- Rotavirus vaccine coverage
- Rubella vaccine coverage
- Sanitation quality
- Second-dose coverage of measles-containing vaccine (MCV2)
- Sex
- Stunting prevalence
- Surrounding conflict
- Time
- Time to care seeking
- Total fertility rate
- Travel time to major city or settlement
- Travel time to nearest health care facility
- Tuberculosis prevalence
- Under-five mortality rate
- Underweight prevalence
- Vaccination efficacy
- Vaccination schedule
- Vaccine coverage equity
- Vitamin A deficiency prevalence
- Vitamin A supplementation prevalence
- Vitamin A treatment prevalence
- Wasting prevalence
- Water quality
- Yellow fever vaccination coverage

### Section 3. Post-discussion list of identified indicators related to measles case fatality

- Access to ICU
- Age
- Ambient air pollution
- Antibiotic use for measles-related pneumonia
- Average household size
- Breastfeeding prevalence
- De-worming frequency
- Diarrheal disease prevalence
- Educational attainment
- Health expenditure per capita
- HIV prevalence
- HIV treatment prevalence / ART prevalence
- Level of health care available
- LRI prevalence
- Malaria prevalence
- Maternal antibody dynamics
- Maternal measles vaccination coverage
- Measles attack rate / incidence
- MCV1 coverage
- MCV2 coverage
- ORT/S for measles-related diarrhea
- Outbreak setting indicator
- PCV vaccine coverage
- Percent living in urban setting
- Population density
- Pre-term birth prevalence
- Sanitation quality
- Surrounding conflict
- Stunting prevalence
- Time to care seeking
- Total fertility rate
- Travel time to major city or settlement
- Travel time to nearest health care facility
- Under-five mortality rate
- Underweight prevalence
- Vaccine coverage equity
- Vaccination efficacy
- Vaccination schedule
- Vitamin A deficiency prevalence
- Vitamin A supplementation
- Vitamin A treatment
- Wasting prevalence

### Section 4. Ranked list of indicators related to measles case fatality, with average rank.

1. Age (1.86)
2. MCV1 coverage (8.43)
3. Underweight prevalence (9.71)
4. Wasting prevalence (9.71)
5. Vitamin A treatment (13.29)
6. Travel time to nearest health facility (13.86)
7. MCV2 coverage (14.00)
8. Level of health care available (14.57)
9. ORT/S for measles-related diarrhea (14.86)
10. Antibiotic use for measles-related pneumonia (15.14)
11. Time to care seeking (15.71)
12. Vitamin A deficiency prevalence (15.71)
13. Stunting prevalence (18.00)
14. Measles incidence (19.57)
15. Surrounding conflict (20.00)
16. Health expenditure per capita (20.14)
17. Access to ICU (20.29)
18. Measles attack rate (20.43)
19. Under-5 mortality (20.71)
20. LRI prevalence (21.00)
21. Diarrheal disease prevalence (21.29)
22. Average household size (21.71)
23. Outbreak setting indicator (23.00)
24. Vitamin A supplementation (23.29)
25. HIV prevalence (23.71)
26. Travel time to nearest city or settlement (23.71)
27. Population density (25.43)
28. Sanitation quality (25.43)
29. HIV treatment prevalence / ART prevalence (25.86)
30. Maternal (measles) vaccination coverage (26.00)
31. Preterm birth prevalence (26.00)
32. TFR / average children per woman (26.86)
33. Percent living in urban setting (27.00)
34. Proxy for maternal antibody dynamics (27.71)
35. Proxy for vaccine coverage equity (28.71)
36. PCV vaccine coverage (30.29)
37. Educational attainment (30.71)
38. Vaccination efficacy (32.14)
39. Ambient air pollution (32.71)
40. Malaria prevalence (33.71)
41. De-worming frequency (34.43)
42. Vaccination schedule (36.71)

### Section 5. PRISMA checklists

#### Section 5a. PRISMA compliance checklist

| Section and Topic | Item # | Checklist item | Location where item is reported |
|---|---|---|---|
| **TITLE** | | | |
| Title | 1 | Identify the report as a systematic review. | Title; identified as literature review |
| **ABSTRACT** | | | |
| Abstract | 2 | See the PRISMA 2020 for Abstracts checklist. | Supplementary Information Section 5b |
| **INTRODUCTION** | | | |
| Rationale | 3 | Describe the rationale for the review in the context of existing knowledge. | Introduction |
| Objectives | 4 | Provide an explicit statement of the objective(s) or question(s) the review addresses. | Introduction |
| **METHODS** | | | |
| Eligibility criteria | 5 | Specify the inclusion and exclusion criteria for the review and how studies were grouped for the syntheses. | Methods (Literature Review subsection) |
| Information sources | 6 | Specify all databases, registers, websites, organisations, reference lists and other sources searched or consulted to identify studies. Specify the date when each source was last searched or consulted. | Methods (Literature Review subsection) |
| Search strategy | 7 | Present the full search strategies for all databases, registers and websites, including any filters and limits used. | Methods (Literature Review subsection); Supplementary Information Section 6 |
| Selection process | 8 | Specify the methods used to decide whether a study met the inclusion criteria of the review, including how many reviewers screened each record and each report retrieved, whether they worked independently, and if applicable, details of automation tools used in the process. | Methods (Literature Review subsection); Contributions |
| Data collection process | 9 | Specify the methods used to collect data from reports, including how many reviewers collected data from each report, whether they worked independently, any processes for obtaining or confirming data from study investigators, and if applicable, details of automation tools used in the process. | Methods (Literature Review subsection); Contributions |
| Data items | 10a | List and define all outcomes for which data were sought. Specify whether all results that were compatible with each outcome domain in each study were sought (e.g. for | Methods (Literature Review subsection) |

| Section and Topic | Item # | Checklist item | Location where item is reported |
|---|---|---|---|
| | | all measures, time points, analyses), and if not, the methods used to decide which results to collect. | |
| | 10b | List and define all other variables for which data were sought (e.g. participant and intervention characteristics, funding sources). Describe any assumptions made about any missing or unclear information. | Methods (Literature Review subsection) |
| Study risk of bias assessment | 11 | Specify the methods used to assess risk of bias in the included studies, including details of the tool(s) used, how many reviewers assessed each study and whether they worked independently, and if applicable, details of automation tools used in the process. | Contributions |
| Effect measures | 12 | Specify for each outcome the effect measure(s) (e.g. risk ratio, mean difference) used in the synthesis or presentation of results. | Results |
| Synthesis methods | 13a | Describe the processes used to decide which studies were eligible for each synthesis (e.g. tabulating the study intervention characteristics and comparing against the planned groups for each synthesis (item #5)). | N/A as no synthesis was performed |
| | 13b | Describe any methods required to prepare the data for presentation or synthesis, such as handling of missing summary statistics, or data conversions. | N/A as no synthesis was performed |
| | 13c | Describe any methods used to tabulate or visually display results of individual studies and syntheses. | N/A as no synthesis was performed |
| | 13d | Describe any methods used to synthesize results and provide a rationale for the choice(s). If meta-analysis was performed, describe the model(s), method(s) to identify the presence and extent of statistical heterogeneity, and software package(s) used. | N/A as no synthesis was performed |
| | 13e | Describe any methods used to explore possible causes of heterogeneity among study results (e.g. subgroup analysis, meta-regression). | N/A as no synthesis was performed |
| | 13f | Describe any sensitivity analyses conducted to assess robustness of the synthesized results. | N/A as no sensitivity analyses were conducted |
| Reporting bias assessment | 14 | Describe any methods used to assess risk of bias due to missing results in a synthesis (arising from reporting biases). | N/A as no synthesis was performed |
| Certainty assessment | 15 | Describe any methods used to assess certainty (or confidence) in the body of evidence for an outcome. | N/A as no synthesis was performed |
| **RESULTS** | | | |
| Study | 16a | Describe the results of the search and selection process, | Results; Figure 2 |

| Section and Topic | Item # | Checklist item | Location where item is reported |
|---|---|---|---|
| selection | | from the number of records identified in the search to the number of studies included in the review, ideally using a flow diagram. | |
| | 16b | Cite studies that might appear to meet the inclusion criteria, but which were excluded, and explain why they were excluded. | N/A; no studies met this criteria |
| Study characteristics | 17 | Cite each included study and present its characteristics. | Results; Supplementary Information Sections 7-8 |
| Risk of bias in studies | 18 | Present assessments of risk of bias for each included study. | Results |
| Results of individual studies | 19 | For all outcomes, present, for each study: (a) summary statistics for each group (where appropriate) and (b) an effect estimate and its precision (e.g. confidence/credible interval), ideally using structured tables or plots. | Results |
| Results of syntheses | 20a | For each synthesis, briefly summarise the characteristics and risk of bias among contributing studies. | N/A as no synthesis was performed |
| | 20b | Present results of all statistical syntheses conducted. If meta-analysis was done, present for each the summary estimate and its precision (e.g. confidence/credible interval) and measures of statistical heterogeneity. If comparing groups, describe the direction of the effect. | N/A as no statistical analysis was conducted |
| | 20c | Present results of all investigations of possible causes of heterogeneity among study results. | N/A as no synthesis was performed |
| | 20d | Present results of all sensitivity analyses conducted to assess the robustness of the synthesized results. | N/A as no sensitivity analyses were conducted |
| Reporting biases | 21 | Present assessments of risk of bias due to missing results (arising from reporting biases) for each synthesis assessed. | N/A as no synthesis was performed |
| Certainty of evidence | 22 | Present assessments of certainty (or confidence) in the body of evidence for each outcome assessed. | N/A as no synthesis was performed |
| **DISCUSSION** | | | |
| Discussion | 23a | Provide a general interpretation of the results in the context of other evidence. | Discussion |
| | 23b | Discuss any limitations of the evidence included in the review. | Discussion |
| | 23c | Discuss any limitations of the review processes used. | Discussion |
| | 23d | Discuss implications of the results for practice, policy, | Discussion |

| Section and Topic | Item # | Checklist item | Location where item is reported |
|---|---|---|---|
| | | and future research. | |
| OTHER INFORMATION | | | |
| Registration and protocol | 24a | Provide registration information for the review, including register name and registration number, or state that the review was not registered. | This review was not registered |
| | 24b | Indicate where the review protocol can be accessed, or state that a protocol was not prepared. | A protocol was not prepared |
| | 24c | Describe and explain any amendments to information provided at registration or in the protocol. | N/A |
| Support | 25 | Describe sources of financial or non-financial support for the review, and the role of the funders or sponsors in the review. | Declarations of Interest; Contributions |
| Competing interests | 26 | Declare any competing interests of review authors. | Declarations of Interest |
| Availability of data, code and other materials | 27 | Report which of the following are publicly available and where they can be found: template data collection forms; data extracted from included studies; data used for all analyses; analytic code; any other materials used in the review. | Results; Supplementary Information Sections 7-8 |

### Section 5b. PRISMA compliance abstract checklist

| Section and Topic | Item # | Checklist item | Reported (Yes/No) |
|---|---|---|---|
| **TITLE** | | | |
| Title | 1 | Identify the report as a systematic review. | Title; identified as literature review |
| **BACKGROUND** | | | |
| Objectives | 2 | Provide an explicit statement of the main objective(s) or question(s) the review addresses. | Introduction subsection |
| **METHODS** | | | |
| Eligibility criteria | 3 | Specify the inclusion and exclusion criteria for the review. | Methods subsection |
| Information sources | 4 | Specify the information sources (e.g. databases, registers) used to identify studies and the date when each was last searched. | Methods subsection |
| Risk of bias | 5 | Specify the methods used to assess risk of bias in the included studies. | N/A |
| Synthesis of results | 6 | Specify the methods used to present and synthesise results. | N/A |
| **RESULTS** | | | |
| Included studies | 7 | Give the total number of included studies and participants and summarise relevant characteristics of studies. | Results subsection |
| Synthesis of results | 8 | Present results for main outcomes, preferably indicating the number of included studies and participants for each. If meta-analysis was done, report the summary estimate and confidence/credible interval. If comparing groups, indicate the direction of the effect (i.e. which group is favoured). | Results subsection |
| **DISCUSSION** | | | |
| Limitations of evidence | 9 | Provide a brief summary of the limitations of the evidence included in the review (e.g. study risk of bias, inconsistency and imprecision). | Conclusion subsection |
| Interpretation | 10 | Provide a general interpretation of the results and important implications. | Conclusion subsection |
| **OTHER** | | | |
| Funding | 11 | Specify the primary source of funding for the review. | Declarations of Interest |
| Registration | 12 | Provide the register name and registration number. | This review was not registered. |

### Section 6. Full list of indicator-specific search terms for systematic review

("educational attainment" OR "education" OR "educat*" OR "school"
OR "urban" OR "crowding" OR "dens*"
OR "conflict" OR "unrest" OR "war" OR "disobedience" OR "state of emergency" OR "pariah state"
OR "travel to health facility" OR "distance to health facility"
OR "care seeking" OR "care-seeking"
OR "proximity to city" OR "travel time to city" OR "distance to city"
OR "stunting" OR "malnourished" OR "malnutrition"
OR "underweight"
OR "vitamin A supplementation"
OR "vitamin A deficien*"
OR "wasting"
OR "ICU" OR "intensive care"
OR "health expenditure" OR "health spending" OR "spending" OR "healthcare per capita"
OR "health care quality" OR "healthcare quality" OR "health care access" OR "healthcare access"
OR "under 5 mortality" OR "under-5 mortality" OR "under five mortality" OR "under-five mortality" OR "infant mortality" OR "child mortality" OR "under 5 death" OR "under-5 death" OR "under five death" OR "under-five death"
OR "air pollution" OR "smog"
OR "antibiotic" OR "pneumonia"
OR "household"
OR "de-worming" OR "deworming"
OR "diarrhea" OR "rotavirus"
OR "HIV" OR "human immunodeficiency virus" OR "acquired immunodeficiency syndrome" OR "AIDS"
OR "antiretroviral therapy" OR "ART"
OR "malaria" OR "plasmodium falciparum" OR "plasmodium vivax"
OR "lower respiratory infection" OR "LRI"
OR "oral rehydration"
OR "pneumococcal vaccine" OR "pneumococcal conjugate vaccine"
OR "pre-term birth" OR "preterm birth" OR "low birthweight" OR "low birth weight"
OR "total fertility rate" OR "average children per women" OR "average number of children per woman"
OR "births per woman" OR "parity"
OR "measles attack rate" OR "measles transmission"
OR "measles incidence"
OR "maternal antibody"
OR "maternal measles vaccination" OR "maternal measles immunity" OR "maternal measles vaccine"
OR (("second dose" OR "MCV2" OR "first dose" OR "MCV1" OR "vaccination" OR "vaccine" OR "immunization") AND "coverage")
OR "equity"
OR "vaccination efficacy" OR "immunization efficacy"
OR "vaccination schedule" OR "immunization schedule" OR "recommended age of vaccination" OR "recommended age of immunization" OR "dosing schedule"
OR "vitamin A treatment"
)
AND "measles"
AND ("case fatality" OR "CFR" OR "fatality" OR "mortality" OR "morbid*" OR "comorbid*" OR "sever*" OR "complicat*" OR "risk" OR "secondary outcome" OR "death")

### Section 7. Studies containing evidence of an association between measles CFR and specified indicator

| Lead Author | Title | Country | Publication Year | Indicator(s) |
|---|---|---|---|---|
| Aaby, P. | Overcrowding and intensive exposure as determinants of measles mortality | Guinea-Bissau | 1984 | Average household size |
| Aaby, P. | Measles mortality, state of nutrition, and family structure: a community study from Guinea-Bissau | Guinea-Bissau | 1983 | Average household size |
| Aaby, P. | The survival benefit of measles immunization may not be explained entirely by the prevention of measles disease: a community study from rural Bangladesh | Bangladesh | 2003 | MCV1 coverage |
| Ahmed, P. A. | Review of childhood measles admissions at the National Hospital, Abuja | Nigeria | 2010 | Underweight prevalence |
| Alwar, A. J. | The effect of protein energy malnutrition on morbidity and mortality due to measles at Kenyatta National Hospital, Nairobi (Kenya) | Kenya | 1992 | Malnutrition |
| Aurangzeb, B. | Clinical outcome in children hospitalized with complicated measles | Pakistan | 2005 | MCV1 coverage |
| Aurangzeb, B. | Risk factors for mortality among admitted children with complications of measles in Pakistan: An observational study | Pakistan | 2021 | Stunting prevalence, underweight prevalence, MCV2 coverage |
| Avila-Figueroa, C. | [Complications in children with measles] | Mexico | 1990 | Malnutrition |
| Barclay, A. J. | Vitamin A supplements and mortality related to measles: a randomised clinical trial | Tanzania | 1987 | Underweight prevalence |
| Bhuiya, A. | Measles case fatality among the under-fives: a multivariate analysis of risk factors in a rural area of Bangladesh | Bangladesh | 1987 | Educational attainment |
| Burström, B. | Child mortality in Stockholm during 1885-1910: the impact of household size and number of children in the family on the risk of death from measles | Sweden | 1999 | Average household size |
| Choudhry, V. P. | Effect of protein energy malnutrition on the immediate outcome of measles | Afghanistan | 1987 | Malnutrition |

| | | | | |
|---|---|---|---|---|
| Clemens, J. D. | Measles vaccination and childhood mortality in rural Bangladesh | Bangladesh | 1988 | Educational attainment |
| Coetzee, S. | Measles in a South African paediatric intensive care unit: again! | South Africa | 2014 | Underweight prevalence, HIV prevalence |
| Commey, J. O. | Measles in Ghana--1973-1982 | Ghana | 1984 | Malnutrition |
| Courtright, P. | Abnormal vitamin A cytology and mortality in infants aged 9 months and less with measles | Malawi | 2002 | Vitamin A deficiency prevalence |
| Dollimore, N. | Measles incidence, case fatality, and delayed mortality in children with or without vitamin A supplementation in rural Ghana | Ghana | 1997 | Underweight prevalence, MCV1 coverage |
| Dzeyie, K. A. | Measles outbreak investigation at Indo-Myanmar border, Longding District, Arunachal Pradesh, India, 2017 | India | 2021 | Vitamin A treatment |
| Fetuga, M. B. | A ten-year study of measles admissions in a Nigerian teaching hospital | Nigeria | 2007 | Malnutrition, MCV1 coverage |
| Gignoux, E. | Risk factors for measles mortality and the importance of decentralized case management during an unusually large measles epidemic in eastern Democratic Republic of Congo in 2013 | Democratic Republic of Congo | 2018 | Travel time to nearest health care facility, MCV1 coverage, MCV2 coverage |
| Gutu, M. A. | Epidemiology of measles in Oromia region, Ethiopia, 2007-2016 | Ethiopia | 2020 | MCV1 coverage |
| Hussey, G. D. | A randomized, controlled trial of vitamin A in children with severe measles | South Africa | 1990 | Vitamin A treatment |
| Hussey, G. D. | Routine high-dose vitamin A therapy for children hospitalized with measles | South Africa | 1993 | Vitamin A treatment |
| Jeena, P. M. | Infectious diseases at the paediatric isolation units of Clairwood and King Edward VIII Hospitals, Durban. Trends in admission and mortality rates (1985-1996) and the early impact of HIV (1994-1996) | South Africa | 1998 | HIV prevalence |
| Joshi, A. B. | Measles deaths in Nepal: estimating the national case-fatality ratio | Nepal | 2009 | Surrounding conflict, stunting prevalence, MCV1 coverage, vitamin A treatment |
| Lagunju, I. A. | Measles in Ibadan: a continuous scourge | Nigeria | 2005 | Underweight prevalence |

| | | | | |
|---|---|---|---|---|
| le Roux, D. M. | South African measles outbreak 2009 - 2010 as experienced by a paediatric hospital | South Africa | 2012 | Malnutrition, HIV prevalence |
| Lee, C. T. | Increase in Infant Measles Deaths During a Nationwide Measles Outbreak-Mongolia, 2015-2016 | Mongolia | 2019 | Travel time to nearest city or settlement |
| Madhulika | Vitamin A supplementation in post-measles complications | India | 1994 | Malnutrition |
| Malina, R. M. | Epidemiologic transition in an isolated indigenous community in the Valley of Oaxaca, Mexico | Mexico | 2008 | Under-5 mortality rate |
| Meteke, S. | Delivering infectious disease interventions to women and children in conflict settings: a systematic review | Various | 2020 | Surrounding conflict |
| Mgone, J. M. | Control measures and the outcome of the measles epidemic of 1999 in the Eastern Highlands Province | Papua New Guinea | 2000 | MCV1 coverage |
| Moss, W. J. | Measles still has a devastating impact in unvaccinated populations | Various | 2007 | Surrounding conflict, malnutrition, vitamin A deficiency prevalence, MCV1 coverage |
| Moss, W. J. | HIV type 1 infection is a risk factor for mortality in hospitalized Zambian children with measles | Zambia | 2008 | Educational attainment, HIV prevalence |
| Moss, W. J. | Measles | Various | 2017 | Malnutrition, MCV1 coverage |
| Murhekar, M. V. | Measles case fatality rate in Bihar, India, 2011-12 | India | 2014 | Educational attainment, vitamin A treatment |
| Nandy, R. | Case-fatality rate during a measles outbreak in eastern Niger in 2003 | Niger | 2006 | Average household size |
| Nayir, T. | Effects of immunization program on morbidity and mortality rates of vaccine-preventable diseases in Turkey | Turkey | 2020 | MCV coverage |
| Ndikuyeze, A. | Priorities in global measles control: report of an outbreak in N'Djamena, Chad | Chad | 1995 | Educational attainment |
| Nojilana, B. | Estimating the burden of disease attributable to vitamin A deficiency in South Africa in 2000 | South Africa | 2007 | Vitamin A deficiency prevalence |
| Oshitani, H. | Measles infection in hospitalized children in Lusaka, Zambia | Zambia | 1995 | MCV coverage/vaccination status |

| | | | | |
|---|---|---|---|---|
| Oshitani, H. | Measles case fatality by sex, vaccination status, and HIV-1 antibody in Zambian children | Zambia | 1996 | HIV prevalence |
| Rey, M. | Impact of measles in France | France | 1983 | Level of health care available |
| Rosero-Bixby, L. | Socioeconomic development, health interventions and mortality decline in Costa Rica | Costa Rica | 1991 | Under-5 mortality rate |
| Salama, P. | Malnutrition, measles, mortality, and the humanitarian response during a famine in Ethiopia | Ethiopia | 2001 | Surrounding conflict, under-5 mortality rate |
| Samb, B. | Decline in measles case fatality ratio after the introduction of measles immunization in rural Senegal | Senegal | 1997 | MCV1 coverage |
| Samsi, T. K. | Risk factors for severe measles | Indonesia | 1992 | Malnutrition |
| Sepúlveda, J. | Improvement of child survival in Mexico: the diagonal approach | Mexico | 2006 | Under-5 mortality rate |
| Spencer, H. C. | Impact on mortality and fertility of a community-based malaria control programme in Saradidi, Kenya | Kenya | 1987 | Under-5 mortality rate |

### Section 8. Studies containing non-significance evidence of an association between measles CFR and specified indicator

| Lead Author | Title | Country | Publication Year | Indicator(s) |
|---|---|---|---|---|
| Aaby, P. | Vaccinated children get milder measles infection: a community study from Guinea-Bissau | Guinea-Bissau | 1986 | MCV1 coverage |
| Aaby, P. | Measles mortality, state of nutrition, and family structure: a community study from Guinea-Bissau | Guinea-Bissau | 1983 | Malnutrition, educational attainment |
| Aaby, P. | Measles incidence, vaccine efficacy, and mortality in two urban African areas with high vaccination coverage | Guinea-Bissau | 1990 | MCV1 coverage |
| Aaby, P. | Overcrowding and intensive exposure as determinants of measles mortality | Guinea-Bissau | 1984 | Average household size, malnutrition |
| Adu, F. D. | Measles outbreak in Ibadan: clinical, serological and virological identification of affected children in selected hospitals | Nigeria | 1997 | MCV1 coverage |
| Ahmed, P. A. | Review of childhood measles admissions at the National Hospital, Abuja | Nigeria | 2010 | MCV1 coverage |
| Ananthakrishnan, S. | Vitamin A and post measles complications | India | 1993 | Vitamin A deficiency prevalence |
| Ariyasriwatana, C. | Severity of measles: a study at the Queen Sirikit National Institute of Child Health | Thailand | 2004 | HIV prevalence |
| Arya, L. S. | Spectrum of complications of measles in Afghanistan: a study of 784 cases | Afghanistan | 1987 | Malnutrition |
| Aurangzeb, B. | Clinical outcome in children hospitalized with complicated measles | Pakistan | 2005 | Malnutrition, proportion living in urban setting |
| Barclay, A. J. | Vitamin A supplements and mortality related to measles: a randomised clinical trial | Tanzania | 1987 | Vitamin A treatment |
| Coakley, K. J. | A review of measles admissions and deaths in the paediatric ward of Goroka Base Hospital during 1989 | Papua New Guinea | 1991 | Preterm birth, vitamin A treatment |

| | | | | |
|---|---|---|---|---|
| Coetzee, S. | Measles in a South African paediatric intensive care unit: again! | South Africa | 2014 | Antibiotic use, vitamin A treatment, PCV coverage |
| Courtright, P. | Abnormal vitamin A cytology and mortality in infants aged 9 months and less with measles | Malawi | 2002 | MCV1 coverage |
| Coutsoudis, A. | Vitamin A supplementation reduces measles morbidity in young African children: a randomized, placebo-controlled, double-blind trial | South Africa | 1991 | Vitamin A treatment |
| Dollimore, N. | Measles incidence, case fatality, and delayed mortality in children with or without vitamin A supplementation in rural Ghana | Ghana | 1997 | Average household size, educational attainment |
| Donadel, M. | Risk factors for measles deaths among children during a Nationwide measles outbreak - Romania, 2016-2018 | Romania | 2021 | Antibiotic use, PCV coverage, preterm birth, vitamin A treatment, MCV1 coverage, malnutrition |
| Fischer, P. R. | Measles in Zaire: 1987 | Democratic Republic of Congo | 1988 | Malnutrition, MCV1 coverage |
| Gignoux, E. | Risk factors for measles mortality and the importance of decentralized case management during an unusually large measles epidemic in eastern Democratic Republic of Congo in 2013 | Democratic Republic of Congo | 2018 | HH size |
| Gutu, M. A. | Epidemiology of measles in Oromia region, Ethiopia, 2007-2016 | Ethiopia | 2020 | Proportion living in urban setting |
| Hull, H. F. | Increased measles mortality in households with multiple cases in the Gambia, 1981 | Gambia | 1988 | Average household size |
| Joshi, A. B. | Measles deaths in Nepal: estimating the national case-fatality ratio | Nepal | 2009 | Average household size |
| Julien, M. | Changing patterns in pediatric mortality, Maputo Central Hospital, Mozambique, 1980-1990 | Mozambique | 1995 | Malaria prevalence |

| | | | | |
|---|---|---|---|---|
| Khoo, A. | Measles--an experience in Sandakan Hospital, Sabah, 1990 | Malaysia | 1994 | Malnutrition |
| Koster, F. T. | Mortality among primary and secondary cases of measles in Bangladesh | Bangladesh | 1988 | Malnutrition |
| Lagunju, I. A. | Measles in Ibadan: a continuous scourge | Nigeria | 2005 | MCV1 coverage |
| le Roux, D. M. | South African measles outbreak 2009 - 2010 as experienced by a paediatric hospital | South Africa | 2012 | MCV1 coverage |
| Lee, C. T. | Increase in Infant Measles Deaths During a Nationwide Measles Outbreak-Mongolia, 2015-2016 | Mongolia | 2019 | Antibiotic use, malnutrition, MCV1 coverage, vitamin A treatment |
| Mafigiri, R. | Risk factors for measles death: Kyegegwa District, western Uganda, February-September, 2015 | Uganda | 2017 | Malnutrition |
| Markowitz, L. E. | Vitamin A levels and mortality among hospitalized measles patients, Kinshasa, Zaire | Congo, Democratic Republic of the (Zaire) | 1989 | Wasting prevalence |
| Moss, W. J. | Prospective study of measles in hospitalized, human immunodeficiency virus (HIV)-infected and HIV-uninfected children in Zambia | Zambia | 2002 | HIV prevalence |
| Munir, M. | Measles and its problems. A clinical analysis of hospitalized patients under 5 years of age | Indonesia | 1982 | Malnutrition |
| Nandy, R. | Case-fatality rate during a measles outbreak in eastern Niger in 2003 | Niger | 2006 | Vitamin A treatment, travel time to nearest health care facility |
| Ogaro, F. O. | Effect of vitamin A on diarrhoeal and respiratory complications of measles | Kenya | 1993 | Vitamin A treatment |
| Sension, M. G. | Measles in hospitalized African children with human immunodeficiency virus | Zaire (DRC) | 1988 | HIV prevalence |
| Smedman, L. | Nutritional status and measles: a community study in Guinea-Bissau | Guinea-Bissau | 1983 | Malnutrition |
